# FETO MATERNAL OUTCOMES OF SINGLETON BREECH DELIVERIES AT MOI TEACHING AND REFERRAL HOSPITAL, ELDORET, KENYA

**DOI:** 10.64898/2026.03.16.26348568

**Authors:** Doreen Momanyi, Godfrey Shichenje Mutakha, Bruce Otieno Semo, Wycliffe Kosgei, Emily Mwaliko

## Abstract

**Background:** Breech presentation which occurs in approximately 3% to 4% of all women at term, is a major concern for both pregnant mothers and their reproductive healthcare providers. This is because it is associated with increased adverse maternal and perinatal outcomes.

**Objective:** To describe the fetal and maternal outcomes of singleton breech deliveries at Moi Teaching and Referral Hospital (MTRH).

**Methods:** This was a cross-sectional descriptive study. The study participants were women with singleton breech deliveries at a gestation of 28 weeks or more. Hospital records indicated that very few breech deliveries occurred at the facility per year. Therefore, a census of all the eligible women with singleton breech deliveries was taken. A semi-structured interviewer-administered questionnaire was used for data collection.

**Results:** There was a total of 11, 748 singleton deliveries at MTRH during the study period (30th August 2019 to 27th August 2020), of which 125 (1.06%) were singleton breech deliveries. Of these, 75 met the eligibility criteria to participate in the study whereby 65 (86.7%) gave birth through emergency caesarean section while 10 (13.3%) had emergency vaginal breech delivery. Most women (50.67%; n=38) delivered at a gestational age of between 38 - 40 weeks and 72 (96%) of the women enrolled had live births. Most (66.70%) newborns weighed 2500 – 3499grammes with 70 (93.3%) newborns having a 5-minute APGAR score of≥7. The majority (85.3%) of the newborns did not have birth complications however, 5 (6.7%) were admitted to the newborn unit due to birth asphyxia while 1 (1.3%) had delayed aftercoming head. The maternal complications noted were second- and third-degree perineal tears (5.3%), post-partum haemorrhage (4.0%) and anaesthetic complications (1.3%).

**Conclusion:** This study noted that despite the MTRH breech delivery protocol recommendation for caesarean section for breech presentation, 13.3% of the women had vaginal breech deliveries. Birth complications (birth asphyxia, NBU admission and delayed aftercoming head) occurred in about 15% of the newborns regardless of the mode of delivery. Furthermore, 40% of these women sustained second- and third-degree perineal tears.

## Introduction

Breech presentation occurs when the foetus lies longitudinally in the uterus with the caudal pole of the foetus occupying the lower uterine segment as the cephalic pole occupies the uterine fundus. Breech presentations account for about 3-4% of all pregnancies at term (1). It is a major predisposing factor for birth complications among newborns. Planned caesarean section has been a safer mode of delivery compared to spontaneous vaginal delivery among women with breech presentation in labour (2). Although the aetiology of breech presentation in pregnancy is not well known, studies have argued that the major predisposing factors for breech presentation include multiple pregnancies, multiparity, prematurity, pelvic abnormalities, placental and uterine abnormalities, polyhydramnios and restricted fetal growth(3).

Breech presentation is the commonest malpresentation in pregnancy that is of great concern for both pregnant mothers and their reproductive healthcare providers. This is because breech deliveries have been associated with increased maternal and perinatal morbidity regardless of the mode of delivery(4). Majority of women in developing countries present to health facilities in labour having not attended any antenatal clinic during pregnancy complicating the diagnosis of breech presentation and subsequent management (5). The best mode of breech delivery has been a controversial issue over the years (6–8). This is because other studies have associated emergency caesarean deliveries with increased mortality and morbidity (7,9). Therefore, there is need for a local study to determine fetal and maternal outcomes of singleton breech deliveries in Western Kenya.

There are controversies surrounding optimal mode of singleton breech deliveries (10,11). Although Moi Teaching and Referral Hospital has guidelines that recommend planned caesarean sections for women with breech presentations, it gives room for emergency caesarean sections and vaginal deliveries. Therefore, there is need for a local study to determine fetal and maternal outcomes based on the mode of delivery among these women. The findings from this study will inform both the care these women receive, and probable mitigation strategies based on the commonly reported outcomes. Furthermore, local studies influence hospital-based management guidelines especially in tertiary hospitals such as MTRH. We described the fetal and maternal outcomes of singleton breech deliveries at MTRH.

## Methodology

This study was conducted at the Riley Mother and Baby unit of Moi Teaching and Referral Hospital (MTRH) in Eldoret, Kenya. We adopted a cross-sectional descriptive study design to describe the proportion of singleton breech deliveries performed at MTRH as well as the resultant fetal and maternal outcomes. Postnatal mothers aged at least 18 years who had delivered singleton foetuses in breech presentation after 28 weeks of gestation either through emergency caesarean section or vaginal delivery at MTRH’s labour ward were enrolled consecutively. Following a written informed consent, questionnaires were administered to the participants and patients’ files were reviewed for additional information. Demographic information such as age, parity, marital status was obtained using an interviewer administered questionnaire while information on previous obstetrical history such as medical conditions, previous vaginal breech deliveries, last delivery and any complication was drawn from the medical records. Descriptive statistical analysis was done by summarizing categorical variables in frequencies with corresponding proportions. The primary fetal outcomes of interest were APGAR score at 5 minutes, admission to NBU, birth asphyxia and perinatal mortality. The APGAR score ≥ 7 was adequate. Maternal outcomes were mode of delivery and birth-related complications such as postpartum haemorrhage, perineal tears and anaesthetic complications. Participants’ characteristics were presented as frequencies with corresponding proportions. To assess the relationship between mode of delivery and fetal outcomes or maternal complications, a Fisher’s exact test was used at a critical value of p≤0.05. Ethical approval to conduct the study was sought from the Institutional Research Ethics Committee (IREC) at the Moi Teaching and Referral Hospital or Moi University School of Medicine (Approval number, FAN: 0003417). A written informed consent was obtained from every participant before enrolment.

## Results

enrolled 75 women majority (74.7%; n=56) of whom were aged 20-35 years. More than half (56%) of the study participants were multiparous, with more than one third (37.3%) of all the study participants having had a previous delivery within two to five years. Furthermore, 70.7% of all the study participants reported a gestation age at birth of 38 weeks or more while only 4 (5.3%) of the women having a history of breech delivery (Table 1).

**Table 1:** Socio-demographic and Reproductive characteristics of study participants (N=75)

| Participant Characteristic | N | % |
| --- | --- | --- |
| **Age (years)** |  |  |
| <20 | 9 | 12.0 |
| 20-35 | 56 | 74.7 |
| >35 | 10 | 13.3 |
| **Parity** |  |  |
| Primiparous | 27 | 36.0 |
| Multiparous | 42 | 56.0 |
| Grandmultiparous | 6 | 8.0 |
| **Year of last delivery** |  |  |
| <2 years | 28 | 37.3 |
| 2-5 years | 28 | 37.3 |
| >5 years | 19 | 25.4 |
| **Gestational age** |  |  |
| 28-34 weeks | 5 | 6.7 |
| 35-37 weeks | 17 | 22.7 |
| 38-40 weeks | 38 | 50.7 |
| >40 weeks | 15 | 20.0 |
| **Previous vaginal breech delivery** |  |  |
| Yes | 4 | 5.3 |
| No | 71 | 94.7 |

Over the study period, 125(1.045 %) women had singleton breech deliveries. The total number of deliveries was 11,957 for a period of one year. Multiple deliveries were: 202 twin and 7 triplet deliveries. There was a total of 11,748 singleton deliveries. Elective breech C/S deliveries were 33 while emergency breech deliveries were 92. Those with commodities were 17 and the participants who were eligible for analysis were 75. This study assessed immediate perinatal outcomes, 5-minute APGAR score and birth complications as the main fetal outcomes of interest. All women who had vaginal breech delivery had a live newborn within 24 hours from the time of delivery, however, 3 (4.6%) of caesarean sections were fresh still births. Similarly, all vaginal deliveries had a 5-minute APGAR score ≥7 similar to 92.3% of caesarean deliveries. Most newborns did not have birth complications, however, there was a delayed after coming head and admission to newborn unit among one and two newborns respectively from vaginal delivery. Among emergency caesarean deliveries, the common birth complications were birth asphyxia and admission to the newborn unit at 7.7% and 4.6% respectively. Mode of delivery was statistically associated (p=0.006) with birth complication, as there was a higher proportion of birth complications among those delivered vaginally (Table 2).

**Table 2:** Fetal outcomes of singleton breech deliveries seen at MTRH.

| Fetal Outcome | Pattern | Vaginal Delivery<br>n (%) | Emergency caesarean<br>Section<br>n (%) | Total | p-value |
| --- | --- | --- | --- | --- | --- |
| Perinatal Outcome | Live | 10 (100) | 62 (95.4) | 72 (96.0) | 0.488 |
|  | FSB | 0 | 3 (4.6) | 3 (4.0) |  |
| 5-minute APGAR score | 1-3 | 0 | 4 (6.2) | 4 (5.3) | 0.662 |
|  | 4-6 | 0 | 1 (1.5) | 1 (1.3) |  |
| | $\geq 7$ | 10 (100) | 60 (92.3) | 70 (93.4) | |
| Birthweight (grammes) | 1500-2499 | 1 (10) | 5 (7.7) | 6 (8.0) | 0.463 |
|  | 2500-3499 | 8 (80) | 42 (64.6) | 50 (66.7) |  |
| | $\geq 3500$ | 1 (10) | 18 (27.7) | 19 (25.3) | |
| Birth Complications | None | 7 (70) | 57 (87.7) | 64 (85.3) | 0.006 |
|  | Delayed after coming head. | 1 (10) | 0 | 1 (1.3) |  |
|  | Birth Asphyxia | 0 | 5 (7.7) | 5 (6.7) |  |
|  | NBU Admission | 2 (20) | 3 (4.6) | 5 (6.7) |  |

Among the women enrolled in this study with singleton breech presentation of their foetuses, 65 (86.70%) of them gave birth through emergency caesarean section while 10 (13.30%) had vaginal deliveries. The common maternal complications noted among vaginal deliveries were perineal tears while in the emergency caesarean deliveries there were anaesthetic complications and post-partum haemorrhage. The mode of delivery was significantly associated (p<0.001) with having a maternal complication (Table 3).

**Table 3:** Maternal outcomes of singleton breech deliveries seen at MTRH.

| Maternal Complication | Vaginal Delivery<br>n(%) | Emergency caesarean<br>Section<br>n(%) | Total | p-value |
| --- | --- | --- | --- | --- |
| None | 6 (60) | 61 (93.9) | 67(89.4) | <0.001 |
| Anaesthetic | 0 | 1 (1.5) | 1(1.3) |  |
| Perineal Tears | 4 (40) | 0 | 4 (5.3) |  |
| PPH | 0 | 3 (4.6) | 3 (4.0) |  |

## Discussion

The proportion of singleton breech deliveries at Moi Teaching and Referral Hospital between 2019 and 2020 was 1.045%. This finding matches that reported in Nigeria (12) and India (13) at 1.7% and 1.3%. The finding of this study is higher than that reported in Zimbabwe at 0.66% (14) which adopted a retrospective study design over one year in 2017 among 8,439 deliveries recorded in the demographic database. This very low proportion of vaginal breech deliveries in Zimbabwe could be attributed to the difference in the eligibility criteria for enrolling women into the two studies. As opposed to this study which cumulated proportion of breech deliveries using both vaginal and caesarean techniques, in Zimbabwe (14) only women who delivered vaginally were included. Differences in study designs have also been attributed to lack of congruence in study findings, as retrospective studies have been attributed to lots of incomplete data hence another reason for the lower proportion of breech deliveries. On the other hand, the current study adopted a cross-sectional study design where no instances of missing data were experienced.

Secondly, the current study’s findings are lower than the global breech deliveries average estimated at 3-4% annually (6,13). The other studies with proportion of breech deliveries that are below the global average were reported in Finland, India and Nepal(4,15,16). In Finland (15), a breech delivery proportion of 2.4% was reported in a ten-year retrospective study between 2005 to 2014 among 585, 580 deliveries. Similarly low proportions of breech deliveries were also reported in studies conducted in India (16) at 2.8% and Nepal at 2.4% (4). These two studies (4,16) were reported recently and conducted in two countries sharing a geographical boundary within the East Asia region. Furthermore, higher numbers of enrolled participants (21,768) and differences in study designs could influence the difference in proportions in the current study and that conducted in Nepal (4).

Other studies have reported proportions of breech deliveries that are within the global average of 3% to 4%(17–21).In India (19), 3% of the 3090 women recruited cross-sectionally had breech deliveries. This was also the case in Pakistan (17) where 100 women were reported to have had vaginal breech deliveries out of 3,977 deliveries seen at a maternal unit in the Bolan Medical Complex Hospital giving a proportion of 3.6%. This study conducted in Pakistan (17) used a case series study design and was conducted over 11 months. This finding mirrors that reported at Wolisso Hospital in Ethiopia where 3.4% of the women had breech deliveries out of 10,214 women enrolled through a cross-sectional study design (18). Although the current and that from Ethiopia (18)adopted the same study design, there was a difference in the number of enrolled pregnant women with breech presentations. In Norway(20) and Nigeria(21), an equal proportion of 3.4% for breech deliveries were reported. Both studies (20,21) adopted retrospective techniques in data collection but enrolled varying number of participants with different eligibility criteria. In Norway, 16, 794 women were enrolled over a ten-year period (2001 to 2011) who had their foetus presenting in a breech manner and all delivered vaginally at the Sorlandet Hospital in Kristians. On the other hand, the 3.4% proportion of breech deliveries reported in Nigeria (21) over a two-and a half year follow-up period was either vaginal or emergency caesarean deliveries.

Majority (70.7%) of the newborns from women enrolled in this study had a gestational age of 38 weeks or more. This finding matches that from Germany (22) and West Indies(23) where more than half (54%) and 82.8% of the newborns had a gestational age of 30-40 weeks respectively. From the findings of a study conducted in Norway (20), the mean gestational age was reported to be 39.4 weeks. However, this finding is higher than that reported in another study conducted in Germany (24) where the mean gestational age was 36.7 (± 1.2) weeks.

This study stratified birth weight into three categories as low birth weight (1500-2499 grammes), normal birth weight (2500-3499 grammes) and large for gestational age (≥3500 grams) at 8.0%, 66.7% and 25.3% respectively. The most frequent (66.7%) birth weight was 2500 to 3499 grammes which is similar to that reported in Ethiopia at 62%(25) and West Indies (23) at 62.6%. Similarly, the mean birth weight reported in studies conducted in Norway(20) and Germany(24) was 3399 grammes and 2577 (±409) grammes respectively. This similarity could be explained by the fact that a majority of these births occurred at term (after thirty-seven complete weeks of gestation).

APGAR score was one of the fetal outcomes that was assessed in this study. It is a standardized, convenient and generally accepted rapid method of assessing the clinical status of a newborn infant at 1 minute, 5 minutes and ten minutes after birth. It reports the state of the infant immediately after birth and the response of the newborn to resuscitation. It was developed by Dr.Virginia in 1952 and it has been adopted for use globally. The APGAR score at 5 minutes predicts the clinical status of the infant better than the 1 minute APGAR score. At 1 minute the newborn has just gone through the stressful process of labour and so it may not indicate the true picture of the clinical state of the newborn. In this study, APGAR score at 5 minutes was reported. A 5 minute APGAR score of less than 7 was considered a low score while a score of 7 or more was considered normal. A5-minute APGAR score was normal (≥7) among nearly all (93.3%) the newborns in this study. This finding is similar to that reported in Ethiopia at 77.7%(25). In Germany, the authors(24) reported a mean 5-minute APGAR score of 9.44 (± 0.9) which matches the findings of this study. Furthermore, the findings are similar to those from Norway (20) and another study conducted in Germany (22) which reported similar 5-minute APGAR score findings. The similarity of the findings from these studies could explained by the fact that a majority of these deliveries occurred through emergency caesarean section. The findings from this study contrasts that reported in Nigeria (Igwegbe et al.,2010) where 50 percent of the newborns had a 5 minute APGAR score of less than seven. The difference could be explained by the use of vaginal mode of breech delivery in a majority of deliveries.

There was a statistically significant association between mode of delivery and birth complications (p=0.006). The birth complications of interest in this study were: delayed after coming head, birth asphyxia and NBU admission. This study reports that 6.7% of all the newborns were admitted to the newborn unit. This finding is close to that reported in Norway at 9%(20)and Germany at 5.6%(22). However, it contrasts the finding from a study conducted in Ethiopia where 25% of the newborns were admitted to NBU(25). This could be explained by the high numbers of birth asphyxia that were reported in Ethiopia(Assefa et al.,2019).

This finding on birth complications being significantly associated with the mode of breech delivery was also witnessed in other studies conducted in different countries(24,26). Although there was no statistical difference reported in the first study conducted in India (26), higher proportions of birth complications (stillbirth and birth asphyxia) were reported among neonates born of women who had undergone emergency caesarean sections. In Austria (24), there was a statistically significant association between mode of delivery and admission to the newborn unit (p<0.001) as a birth complication. In Canada(6), serious morbidity (p=0.003) and perinatal mortality (p=0.01) was significantly associated with the mode of delivery. This current study’s findings on the significant association between mode of delivery and birth complications are consistent with those reported in other countries across the globe which targeted the same group of women and used similar study designs(6,24).

This study reports a fresh still-birth rate of 4.6% among three women who had emergency caesarean sections with no perinatal mortality noted among those who had vaginal deliveries. However, this relationship was not statistically significant. One of the fresh stillbirths was found to have congenital anomalies, while two women had prolonged labour. This finding is in contrast with that reported in Finland (15) where the stillbirth rate in term breech presentation was significantly higher compared to cephalic presentation.

65 (86.70%) of the women enrolled with singleton breech presentation of their foetuses gave birth through emergency caesarean section compared to 10 (13.30%) who had vaginal deliveries. This finding of more emergency caesarean deliveries compared to vaginal deliveries among women with singleton breech presentation is comparable to the findings from other African countries(25,27). In a study conducted in Ethiopia’s Jimma University Medical Center (JUMC) in Addis Ababa, 61.1% of the term foetuses who were presenting in breech position were delivered through emergency caesarean section (Assefa et al.,2019). Similarly, in Nigeria (27) where the authors compared vaginal and emergency caesarean sections among women with singleton breech presentation; most women delivered through emergency caesarean sections. However, in a different study conducted in Oromia Region of Southern Ethiopia (18), there was a significantly higher proportion (82.6%) of vaginal delivery among women with singleton breech presentation. This difference was still witnessed even though the authors (18) excluded preterm deliveries in a mission hospital in the rural region of Ethiopia with resource constrains. Similarly, in other studies (28,29) conducted in countries outside the continent of Africa higher proportions of vaginal deliveries were reported. In Germany (28), it was reported that there were 74.9% singleton vaginal breech deliveries while in Pakistan (29) 65% of the women had vaginal deliveries. The difference could be explained by the different modes of deliveries used in these studies.

This study reports that most (89.33%) of the study participants did not have any complication associated with breech deliveries. This finding corresponds to other studies reporting on singleton breech deliveries(16,30,31). In a study conducted in India (31), 90% of the women who had singleton breech deliveries did not have any complications noted. However, lower proportions of no complications were reported in Bhubaneswar-India (16) and Nigeria (30) at 82.1%. However, the proportionate differences in lack of complications were not markedly different. These low proportions of complications indicate that singleton breech deliveries (majority of them being emergency caesarean sections) are safe to the women diagnosed with this pregnancy presentation.

Anaesthetic complications were assessed in this study. It was reported this complication occurred at a rate of 1.33%. In this study only one participant had anaesthetic complication which was described as difficult induction. Postpartum hemorrhage is defined as the blood loss of more than 500ml following vaginal delivery or more than 1000mls following emergency caesarean delivery that is associated with hemodynamic instability requiring urgent intervention. There were 4% of the women enrolled with post-partum haemorrhage following singleton breech deliveries which was close to that reported in two studies previously conducted in India at 5.1% (16) and 4.8% (32). However, in Nigeria(30), the proportion of post-partum haemorrhage was found to be 1.3% which is much lower than that reported in this study. However, a different study conducted in India reported a much higher proportion of post-partum haemorrhage at 8% (31), a difference attributed to the difference in the eligibility criteria. Differences in study designs, enrolment of varying numbers of study participants, conducting of the studies in different setups and different study durations could directly influence the proportion of participants with the factor of interest.

Perineal tears were assessed in this study. They are defined as lacerations that develop as a result of injuries to the overstretching perineal tissues that occur during childbirth. The perineum is a diamond-shaped area between the vaginal opening anteriorly and the anal opening posteriorly. Perineal tears due to fetal malpresentation are lacerations of skin and other soft tissues that separate the vagina from the anus. During breech delivery, the perineum may be stretched unevenly by irregular pressure from the breech or the limbs causing tears. In this study, all perineal tears were reported among women who had vaginal deliveries. These tears accounted for 40% of all complications reported among women who had vaginal breech deliveries. This study classified the vaginal tears between first to fourth degrees, depending on the extent of the laceration. The first-degree tear is limited to the mucosa and skin of the introitus. The second degree involves the fascia and muscles of the perineum while the third degree involves the anal sphincters. Lastly, fourth-degree tears have the trauma extending to the rectal lumen through the mucosa. Among the women with perineal tears, those with second and third-degree perineal tears required repair. Three women sustained second-degree perineal tears that were repaired in labour ward while one woman had a third-degree perineal tear that was repaired in the theatre. This study collected perineal tears data because of its associated risk for infections, post-partum haemorrhage, disfiguration, and faecal incompetence. However, the findings of this study contrast those reported in a retrospective study conducted in Nepal (4) where no cases of third- and fourth-degree tears were reported. The authors (4) noted that their study design and sample size were not strong enough to provide conclusive findings. Secondly, the authors did not compare the outcomes of vaginal delivery with emergency caesarean delivery. Because of this, they could not infer from the study that the higher rate of perinatal morbidity and mortality can be attributable to vaginal mode of delivery. In another study conducted in two French tertiary care centres of Paris suburbs (33), perineal tears occurred in 63% of the women who delivered macrosomic infants. This difference could be attributed to the scope of the study in France (33), compared to that of the current study. The French study was conducted between 2005 to 2008 among 27,630 patients who delivered in the two tertiary hospitals compared to the current study conducted in a single tertiary hospital among 75 mothers. In advanced labour uncomplicated breech deliveries can be conducted either through emergency caesarean delivery or vaginal delivery because there was good perinatal outcome irrespective of the mode of delivery.

Because maternal complications were associated with the mode of delivery, efforts should be made to ensure that those with breech presentations are identified during antenatal visits, admitted at term and prepared for elective caesarean section. In addition, those with breech presentation in labour should be prepared for emergency caesarean section.

## Conclusions and Recommendations

This study noted that despite the caesarean section recommendation for singleton breech deliveries at MTRH, 13.3% of the women had vaginal breech deliveries. Birth complications including birth asphyxia, admission to the newborn unit, and delayed aftercoming head occurred in < 15% of the newborns regardless of the mode of delivery. Furthermore, 40% of these women sustained second- and third-degree perineal tears. This study contributes to the knowledge on fetal and maternal outcomes of singleton breech deliveries at a national teaching and referral hospital located in Western Kenya. Previous studies have focused on the association between mode of delivery and fetal outcomes; however, this study went ahead to assess both maternal and perinatal outcomes. In advanced labour uncomplicated breech deliveries can be conducted either through emergency caesarean delivery or vaginal delivery because there was good perinatal outcome irrespective of the mode of delivery.

Because maternal complications were associated with the mode of delivery, efforts should be made to ensure that those with breech presentations are identified during antenatal visits, admitted at term and prepared for elective caesarean section. In addition, those with breech presentation in labour should be prepared for emergency caesarean section.

## Data Availability

All data produced in the present study are available upon reasonable request to the authors

## Acknowledgement

We are grateful to the mothers who participated in this study as well as the hospital’s staff and management.

